# The Impact Of COVID-19 on Health Financing in Kenya

**DOI:** 10.1101/2023.04.05.23288177

**Authors:** Angela Kairu, Stacey Orangi, Boniface Mbuthia, Brian Arwah, Fatuma Guleid, Janet Keru, Ileana Vilcu, Anne Musuva, Nirmala Ravishankar, Edwine Barasa

**Affiliations:** Health Economics Research Unit (HERU), KEMRI-Wellcome Trust Research Program, Nairobi, Kenya; ThinkWell Kenya, Nairobi, Kenya; ThinkWell, Regus, Geneva, Switzerland; Thinkwell Washington DC, USA; Centre for Tropical Medicine and Global Health, Nuffield Department of Medicine, University of Oxford, Oxford, United Kingdom

## Abstract

**Background:** Sudden shocks to health systems, such as the COVID-19 pandemic may disrupt health system functions. Health system functions may also influence the health system’s ability to deliver in the face of sudden shocks such as the COVID-19 pandemic. We examined the impact of COVID-19 on the health financing function in Kenya, and how specific health financing arrangements influenced the health systems capacity to deliver services during the COVID-19 pandemic.

**Methods:** We conducted a cross-sectional study in three purposively selected counties in Kenya using a qualitative approach. We collected data using in-depth interviews (n = 56) and relevant document reviews. We interviewed national level health financing stakeholders, county department of health managers, health facility managers and COVID-19 healthcare workers. We analysed data using a framework approach.

**Results:** Purchasing arrangements: COVID-19 services were partially subsidized by the national government, exposing individuals to out-of-pocket costs given the high costs of these services. The National Health Insurance Fund (NHIF) adapted its enhanced scheme’s benefit package targeting formal sector groups to include COVID-19 services but did not make any adaptations to its general scheme targeting the less well-off in society. This had potential equity implications. Public Finance Management (PFM) systems: Nationally, PFM processes were adaptable and partly flexible allowing shorter timelines for budget and procurement processes. At county level, PFM systems were partially flexible with some resource reallocation but maintained centralized purchasing arrangements. The flow of funds to counties and health facilities was delayed and the procurement processes were lengthy. Reproductive and child health services: Domestic and donor funds were reallocated towards the pandemic response resulting in postponement of program activities and affected family planning service delivery. Universal Health Coverage (UHC) plans: Prioritization of UHC related activities was negatively impacted due the shift of focus to the pandemic response. Contrarily the strategic investments in the health sector were found to be a beneficial approach in strengthening the health system.

**Conclusions:** Strengthening health systems to improve their resilience to cope with public health emergencies requires substantial investment of financial and non-financial resources. Health financing arrangements are integral in determining the extent of adaptability, flexibility, and responsiveness of health system to COVID-19 and future pandemics.

## Introduction

The first case of COVID-19 in Kenya was reported on 13^th^ March 2020 soon after the World Health Organization (WHO) declaration. As of 11^th^ August 2022, Kenya had recorded 337,912 infections and 5,673 deaths from COVID-19 (1). So far, the country has experienced five COVID-19 epidemic waves (early August 2020 (wave 1), late-November 2020 (wave 2), mid-April 2021 (wave 3), late August 2021 (wave 4), and mid-January 2022 (wave 5)) at the time of this manuscript preparation (2).

To limit the spread of infection, Kenya adopted several strategies to enable the health system to contain the pandemic and cope with the demand of COVID-19 health services. The non-pharmaceutical interventions (NPIs) implemented included closure of borders, restriction of movement across the country and an international travel ban except for cargo, closure of school/learning institutions, ban on religious and social gatherings and meetings, a dawn to dusk curfew, and social physical distancing (1.5 m) in areas of gathering. The government progressively lifted these restrictions based on the trend of infections over the pandemic period (3). Furthermore, pharmaceutical interventions for treatment of COVID-19 patients were implemented based on the Kenya case management guidelines (4, 5). For sustainable control of the pandemic, the COVID-19 vaccination campaign targeting 1.02 million health workers and those above the age of 58 years was launched on 5^th^ March 2021, and later extended to cover all above 18 years old (6). As of 1^st^ April 2022, 8,090,985 individuals were reported to be fully vaccinated (7).

The interaction between the COVID-19 pandemic and the health system functions is bi-directional. One is the capacity of health system functions which affects the effectiveness of the country’s response to the pandemic, and on the other hand the nature, scale, health and non-health impacts of the pandemic, and country response strategies affect health system functions in ways that influence the resilience of health systems. The health financing functions may influence the health system’s ability to support continued good quality service delivery. Specifically, resilience in revenue collection, risk pooling, purchasing and service delivery is key in the health system’s response to crises (8). Understanding these interactions by evaluating the existing health financing arrangements considering the pandemic is important to strengthen the health system and prepare for future pandemics.

Against this background, the study examined how the COVID-19 pandemic and government response impacted the health financing system, the effect of the existing health financing arrangements on the capacity of the health system to respond to the pandemic, the adaptations made to better the health system’s response to the pandemic, and the influence of the pandemic response on the effectiveness of health financing system to promote health system goals and universal health coverage (UHC). Specifically, we examined the purchasing and public finance management (PFM) dimensions of the health financing system in general, and also used the financing of reproductive, maternal, neonatal, child, and adolescent health (RMNCAH) as a specific tracer for the impact of COVID-19 on health financing.

## Methodology

### Country Context

Kenya is a lower-middle-income country with a GDP per capita of $2,006.80 (2021) (9). The country’s population in 2019 was estimated at 47.56 million people with a predominantly young population (10). In 2013, Kenya transitioned to a devolved system of governance comprising of the national government and 47 semi-autonomous county governments. Under devolution, regulatory and policy functions in health were maintained at a national level while health service delivery functions were transferred to county governments (11). The health sector comprises of both the public and the private sector and is characterized by six levels of health facilities: I) community health units; II) dispensaries; III) health centres; IV) county hospitals; V) county referral hospitals; VI) national referral hospitals (11).

Kenya’s health system is financed through four main sources: government, out of pocket (households), donors, and private-estimated at 45.98%, 24.3%, 18.51%, and 35.51% of total health expenditure in 2018/19 (12). There are three main purchasers in the public health system, as described in Table 1. First, the National Ministry of Health (MoH) pays for health services at public tertiary health facilities through global budgets (13). Second, county governments pay for health services offered at public primary and secondary health facilities through payment of salaries of health care workers, supply of commodities, and line-item budgets (13). Third, the National Health Insurance Fund (NHIF) pays for outpatient services, inpatient services, and maternity packages for schemes to registered NHIF members in public and contracted private and faith-based facilities, and reimburses using different provider payment mechanisms (14, 15). Further, the country’s free maternity program, dubbed *Linda Mama,* is purchased by the NHIF through contracted facilities, across all types and levels and reimbursements done through case-based payments (16). There also are private insurers and community-based health insurance (CBHI) schemes which purchase health services through private contracts with facilities.

**Table 1:** Purchasing in the public health system in Kenya

| Purchasers | Purchasing arrangement | Benefit packages | Type of facilities | Populations covered | Provider payment mechanisms |
| --- | --- | --- | --- | --- | --- |
| National MoH | Public integrated arrangement | Outpatient and inpatient services | Public tertiary facilities | Kenyan citizens | Global budgets |
| County government |  | Outpatient and inpatient services | Public primary and secondary facilities | Kenyan citizens (county residents) | -Line-item budgets<br>-Salaries<br>-Supply of commodities |
| NHIF | Public contract | Outpatient services | Public and contracted private and faith-based facilities | Registered NHIF members | Capitation and fee for service (dependent on scheme) |
|  |  | Inpatient services | Public and contracted private and faith-based facilities | Registered NHIF members | Rebates and bundled payments |
|  |  | Free maternity program and maternity packages for schemes | Public and contracted private and faith-based facilities | Registered NHIF members and those registered for the FMP | Case-based payment |
| Private insurers, CBHI schemes | Private contract | Outpatient and Inpatient services | Public and contracted private and faith-based facilities | Registered members | Agreed upon provider payment method (PPM) with the providers |

### Conceptual framework

We applied a conceptual framework which assumes linkages between health financing functions, health system resilience, and progress in the attainment of broader health system goals and UHC (Fig 1). First, the framework assumes that, in the face of a pandemic, some health financing arrangements may remain unchanged, some may be disrupted, and some may be adapted to enhance the health systems response to the pandemic. This financing arrangements include revenue collection, risk pooling, purchasing, and PFM. Second, the framework assumes that the status (maintained, disrupted, and/or adapted) of health financing arrangements will influence the health systems resilience to the COVID-19 pandemic. The health system resilience refers to the health systems capacity to respond to the pandemic by a) mitigating and containing the pandemic b) maintaining core health system functions and delivery of core health services and c) minimizing disruptions of existing health system plans, policies, and priorities. The resilience of the health system also has the potential to influence the state of health financing functions. Third, the resilience of health systems may influence country progress to attain health system goals and UHC. These include equitable access to quality health services and financial risk protection, underlined by principles of equity, efficiency, accountability, and responsiveness of the health system to citizen needs. In turn, progress towards these goals may influence health system resilience.

**Figure 1:**
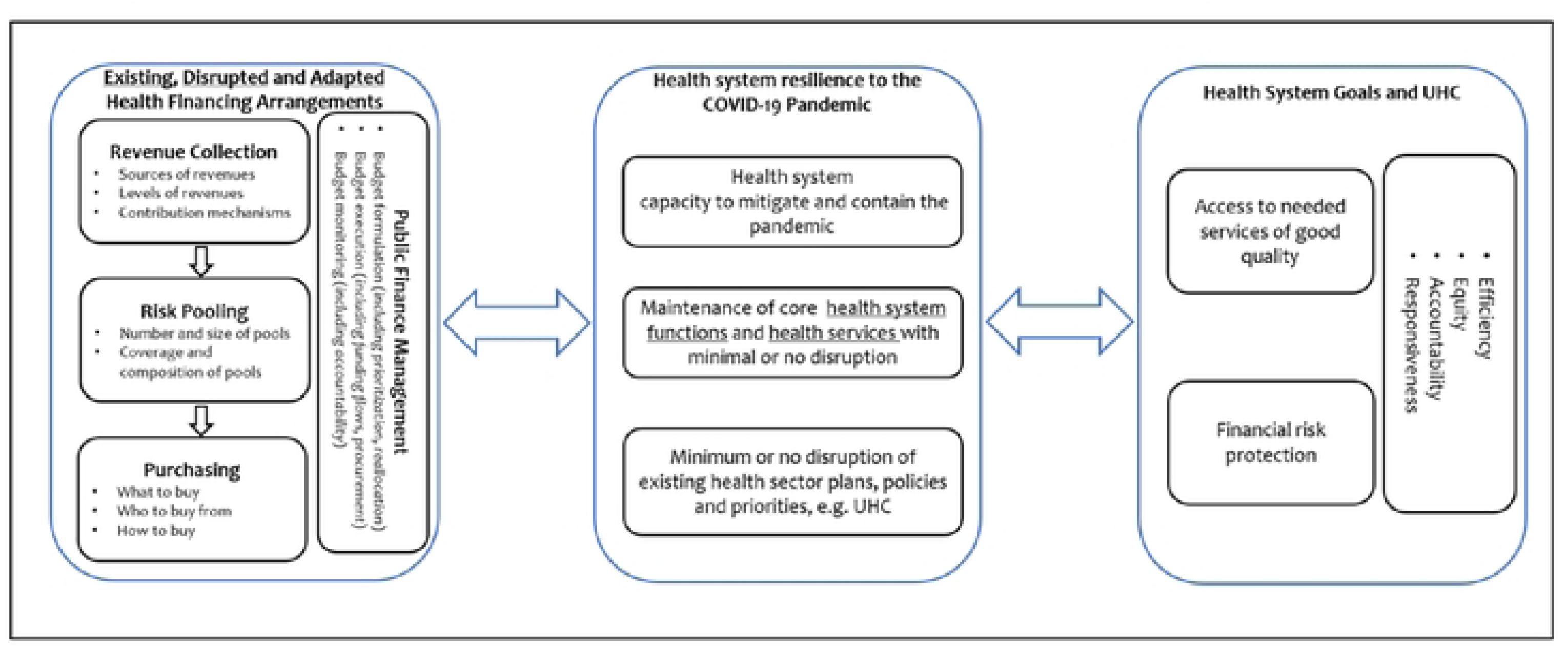
Conceptual framework for the impact of COVID-19 on health financing

### Study design

We conducted a cross sectional study where we employed qualitative methods for data collection. This approach allows for analysis of in-depth individual interviews considering the complexity, detail and context (17).

### Study sites, population, data collection

We purposively sampled three counties, guided by negotiations with the MoH and the Council of Governors to reflect convenience, geographical variation, and variation in health financing arrangements. We have anonymized the counties to maintain confidentiality of the study participants. In each county, we selected six health facilities to represent the different types and levels of service delivery: one county referral (level 5) hospital, one government health centre (level 3), one faith-based hospital (level 4), one faith-based health centre (level 3), one private hospital (level 4) and one private health centre (level 3). Approval to conduct the study in these health facilities was obtained from the different institutional authorities.

We collected data between October and December 2021 through in-depth interviews (IDIs) and document reviews. All study participants were presented with information on the organization conducting the study, the purpose of the study, and who the researchers were, and gave their written informed consent. Four researchers (AK, SO, BA and FG) conducted 56 IDIs in English with participants from the national, county and facility levels (Table 2) using semi-structured interview guides developed in reference to health financing arrangements (See Additional file 1: Semi-structured interview guide). The validity of the semi-structured interview guides was tested by a team of health economic researchers in our research organization and the collaborating institution in Kenya, to check for ambiguities and leading questions. All IDIs were conducted at the participant’s workplace and were audio-recorded using encrypted audio-recorders. Each IDI lasted between 50 and 60 min. Four researchers held face-to-face peer de-briefing sessions after conducting IDIs to critique the data collection process and identify areas that needed further probing (18). We stopped data collection once saturation point of no new information was reached (19).

**Table 2:**
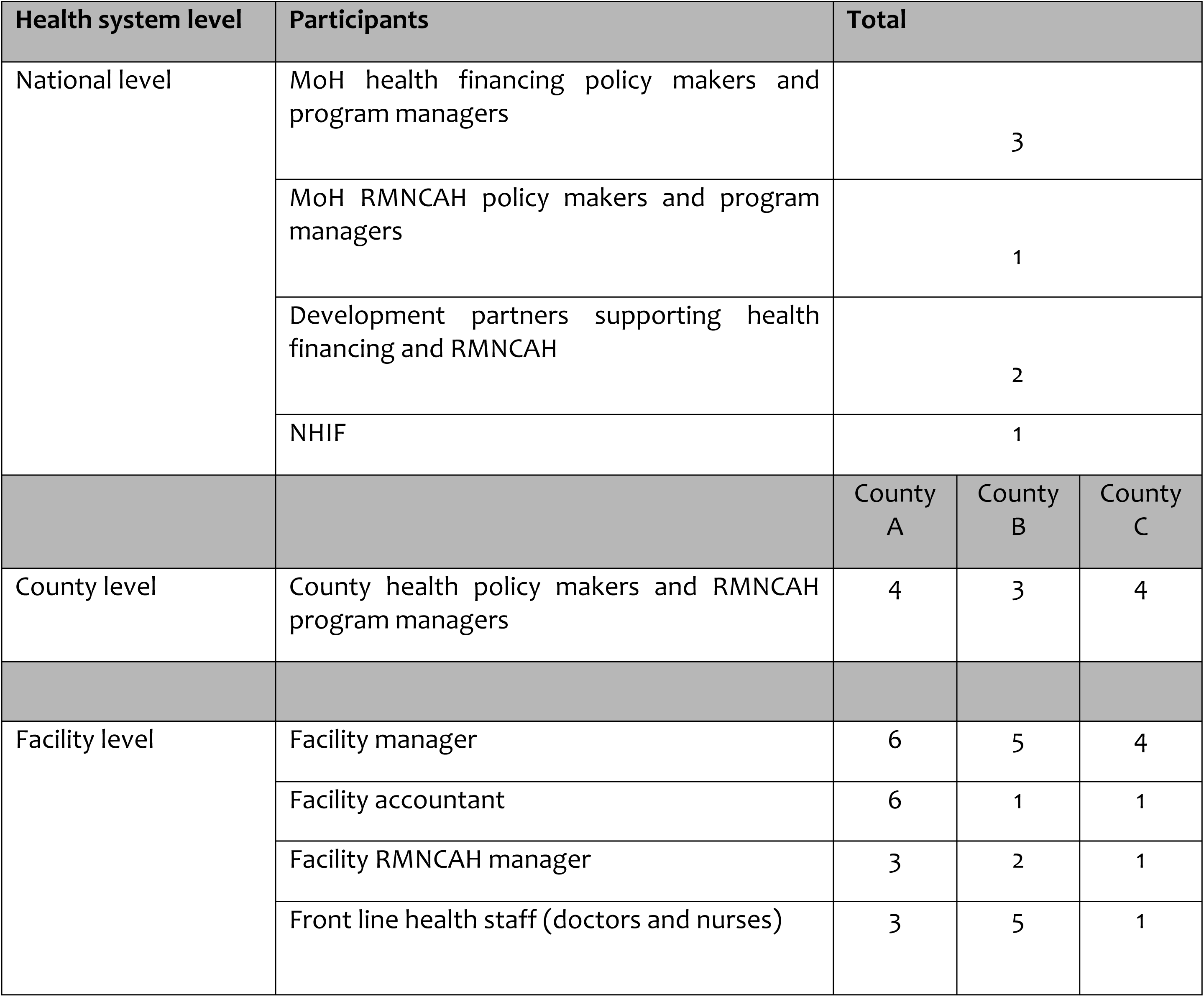
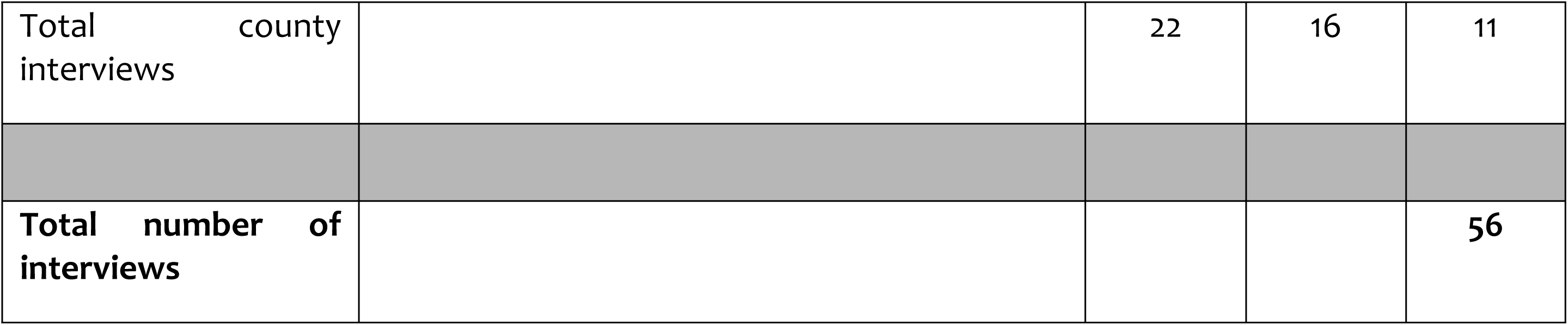
Summary of study participants

### Sampling and sample size

We purposively selected respondents with knowledge of and experience in health financing arrangements which was the phenomenon of interest in our study (Table 2). We selected participants at the national, county, and facility levels. Participants at the national level included health financing stakeholders (policy makers, implementers), and development organizations providing technical support to health financing initiatives in Kenya. At the county level, participants included county department of health officials, health facility managers and administrators and COVID-19 healthcare workers at facility level.

We reviewed published policy documents, press releases, grey and peer reviewed literature regarding current health financing arrangements in Kenya.

### Data management and analysis

All audio records from the IDIs were transcribed verbatim in English. All transcripts were reviewed against their respective audio files for transcription accuracy. The validated transcripts were then imported to NVIVO 10 for coding guided by the topic areas. We used a framework approach to analyse data. This approach involves a process of systematic sifting, sorting, coding, and charting data into key issues and themes (20). One researcher (AK) first familiarized herself with data by reading and re-reading the transcripts. She developed codes deductively from the conceptual framework and applied the codes to interpret segments in the transcripts that were important. The study team members (AK, SO and EB) reviewed and discussed the initial coding framework, and any discrepancies were appropriately reconciled. The final coding framework was applied by (EB and AK) to the data and later charted the data to allow the emergence of themes through comparisons and interpretations.

### Ethical Considerations

This study received ethics approval from the KEMRI Scientific and Ethics Review Unit (SERU), approval number KEMRI/SERU/CGMR-C/132/3735, Kenya, National Commission for Science, Technology and Innovation (NACOSTI) serial no. A17531, Council of Governors prior to data collection. All study participants were presented with information on the organization conducting the study, who the researchers were, the purpose of the study, the right to withdraw and measures put in place to ensure confidentiality and gave their written informed consent. Informed consent both written and oral was obtained from potential participants before the interviews were conducted. Participants were informed that data will be reported in an aggregated format and anonymity will be ensured in storage and publication of the findings of the study.

## Results

### Purchasing Arrangements

#### **i)** What to purchase

In Kenya, the defined set of COVID-19 services offered included COVID-19 testing, isolation, case management (treatment), and vaccination. The range of COVID-19 services provided was determined at the national level through a consultative process. Two main teams, namely the National Emergency Response Committee, and the National Taskforce were established to lead the implementation of response strategies. Technical working groups (TWGs) were established under the National Taskforce to determine the services and develop guidelines based on global experiences, reference to global documents and guidelines and consultation with developmental partners, public-private research, and academia. The clinical management protocol developed was then disseminated countrywide to guide clinical management of COVID-19 patients.

> *“That was a consultative process that drew from public-private research academia, development partners, through various task forces, committees, technical working groups and joint documents were released and published. National official 2, MoH*

***Variation in inclusion of COVID-19 services in the NHIF benefit package promoted inequity in access to COVID-19 services.*** The NHIF adapted its enhanced scheme’s (civil service, national police service, prison service) benefit package to include COVID-19 services through establishment of a separate pool of funds financed by the employer. This fund catered for COVID-19 testing, treatment (in-patient) and consumables related to COVID-19 management. The NHIF did not do a similar adaptation to its general scheme which covers the rest of the population, including those in the informal sector and the poor. This meant that only the formal sector working in government institutions benefited from NHIF insurance cover for COVID-19 services.

> *“A provision was made for enhanced schemes purchased by government employers (public service commission for civil service, the police, and the prison service), which set out additional funds specifically to take care of COVID-19 for their members. This is a separate basket from the premiums. It covers the testing, admission both in the general ward and the ICU, and the consumables that are related to management of COVID-19. It is a pool of funds that is available to members who are covered within that scheme, both the primary or the principal member and the declared beneficiaries” National official 4, NHIF*

***Individuals belonging to the general NHIF scheme or those without health insurance were thus exposed to out-of-pocket expenditure seeking COVID-19 services across the different types of health facilities***. This included sample referral for testing and admission referrals to other counties.

> *“We do not have a cover for Covid-19, so somebody pays out of pocket unless the county government agrees to waive the bill. If you are a member of NHIF, and you’ve been taken into isolation for treatment, then you pay from your pocket. But we cover a group of people, the National Police Service, includes the Prisons and everybody under the National Scheme” NHIF staff, County C*

> *“Patients pay out of pocket, if it’s not the NHIF enhanced schemes, if testing is done elsewhere” Facility administrator, private facility, County A*

Additionally, there was limited coverage for COVID-19 services from private health insurance institutions specific to accredited public hospitals. Therefore, individuals with private health insurance were also exposed to out-of-pocket expenditure.

> *“Within this facility you find that most insurances (private insurance) don’t cover for the COVID 19 patients, so they pay for themselves.” Nurse In-charge, private facility, County B*

For the individuals unable to afford COVID-19 services, the county governments absorbed these costs in two ways. First, the county governments conducted contact tracing activities, and supplied health facilities with PPEs, testing reagents and kits and vaccines, to subsidize the costs of testing in private facilities and provision of free vaccination services in both public and private health facilities.

> *“Most of the health services related to COVID-19 response and management are free within the county. The contact tracing, testing, vaccination. The costs are catered for by the county government. Also, the county government procures the commodities to be used” County Chief Officer of Medical Services, County A*

> *“The county provides the test kits, but any other services apart from providing the test kits are borne by the facility. In terms of treatment and isolation, our facility bears the cost, which is then transferred to the patient. The facility offers vaccination, which is not paid for. It’s free.” Clinical Officer, FBO hospital 1, County B*

Second, pending hospital bills for COVID-19 admissions in public hospitals were waived and paid by the county using the allocated COVID-19 funds, and additionally for County B through the county universal healthcare scheme. However, medication and diagnostic services were limited to the list of the essential medicines and diagnostics on the healthcare scheme. This implied further out-of-pocket expenditure incurred by individuals.

> *“A patient with no insurance card like the UHC or NHIF pays for services in cash. If you have these insurances, you don’t pay for anything. Every service that is in the facility is covered by the UHC card except for services like the imaging or the more expensive drugs that are not available. You must buy or maybe pay for imaging service elsewhere.” Nurse COVID ward, hospital 1, County B*

> *“Currently, the testing is free. Then on isolation and treatment, the same is waived by the county government. However, if the diagnostic test and medication is outside the insurance essential list, the patient pays. Yes.” County Director of Health Planning, County B*

There were various challenges experienced with availability of COVID-19 healthcare services paid for by the government. At the national level these included delays in procurement due to shortage of commodities (vaccines, supplies) attributed to global shortages, lengthy logistics (from manufacturers, suppliers and to target countries), high prices of oxygen and high demand that almost out-stripped supply, and scarce in-country financial resources. For the counties, there was inadequate financial resources to deliver services, stock out of commodities (testing kits, reagents, PPEs, medications, vaccines), and high costs of processing test samples in national laboratories.

A mixed approach was adopted to include centralised national procurement process (for essential supplies laboratory, medicines and non-pharmaceuticals). This approach combined resources from national government and donors through KEMSA, direct procurements by counties through KEMSA and from other suppliers, and, direct supplies through implementing partners (private and NGOs) that were provided directly to identified high burden COVID-19 counties.

#### **ii)** From whom to purchase

While both private and public facilities provided COVID-19 services, public health facilities provided a larger share of these services. The national government progressively prioritized direct support to health facilities in different counties as the disease burden increased. The MoH through the multi-agency teams accredited health facilities progressively as they determined their capacity to offer COVID-19 services. The list of accredited facilities and isolation/quarantine centers was made available to the public. The national level laboratories and private laboratories listed by MOH were the main COVID-19 testing centres for samples countrywide. Public and private level 4-6 hospitals provided testing, treatment, isolation, and vaccination services, whereas level 2 and 3 were only used as quarantine centres at the start of the pandemic and later reverted to routine health services. This implied that a significant proportion of individuals accessed the services in government facilities.

> *“In our facility, we are only offering screening and vaccination. In case we get cases, we suspect for COVID-19, we refer to the sub-county hospital” Nurse in charge, public health centre 1, County B*

> *“We have relied on the publication by the ministry on providers who have been evaluated by the health care systems or establishments at the ministry that include the medical council, Kenya Health Professions Oversight Authority (KHPOA), and have been certified as being able to offer COVID-19 services. So those are the facilities that we have engaged, but it’s dependent on the line lists that are provided by other government agencies.” National official 4, NHIF*

During infection surges, government hospitals had inadequate capacity to provide COVID-19 services characterized by limited hospital bed capacity, healthcare workers, supplies (PPEs) and ambulances for referral of patients.

> *“There are challenges in the provision of PPEs, referral of patients, cost of training staff because the facilities must pay for these. The cost of attending to these clients is high, which means that the facilities must adjust their budgets to factor in the COVID-19.” Clinical Officer, FBO hospital 1, County B*

> *“At times we have one ambulance, and it is the same ambulance that we are using in the whole sub county also for maternity cases. Also, it’s the general nurses and health workers who attend to the COVID-19 patients because of shortage of staff.” Nursing officer in-charge, public hospital 2, County C*

Only a few private health facilities provided COVID-19 services within the study counties. These services were paid for either by the NHIF enhanced scheme or by individuals as out-of-pocket expenses. This excluded vaccination services paid for by the national government. Only insured individuals or those who were able to afford the costs accessed services in private health facilities.

> *“The county residents can get the vaccines from private facilities for free which are being supplied by the county department. But there is no payment done by the county for other COVID-19 services to private facilities.” County Official 2, County B*

> *“If the residents choose to seek services from the private facilities, then they pay for them apart from the services covered by national government.” National official 2, MoH*

In addition, access of services in the private sector was limited by some private health facilities declining to admit patients without COVID-19 test results in county C, and inadequate COVID-19 ward bed capacity resulting in transfer of patients to government facilities.

> *“For a good number of private facilities, it’s a requirement to have a COVID-19 test before admission. The chances of being turned away is very high.” County Nurse In charge, County C*

> *“The commonest challenge would be either the numbers and the capability of the private hospitals not able to take care of the patients and also versus our numbers because we may find also there are situations where our CTU is packed and the private also has patients they want to bring over.” Maternity nurse in-charge, public hospital 1, County B*

***The county governments put in place certain arrangements for county residents to seek COVID-19 services in private facilities***. First, the counties informally engaged private health facilities for referral of patients from government facilities. Second, the government provided private facilities with select COVID-19 testing supplies and vaccines to subsidize the costs in the facilities.

> *“The private hospitals are given the vaccines by the county and the citizens can walk into any facility get it for free. But for the other curative services, they must pay because now those are private entities.” COVID-19 coordinator, County B*

> *“For testing, private facilities are provided with the test kits by the county government so that they do not charge the patients, although I know probably some of the ones which acquire their own test kits from elsewhere apart from the county stores, they charge for testing. For vaccination, the government agreed with the private and faith-based hospitals to list them as vaccination centers if they meet the requirements for vaccination.” COVID-19 coordinator, County A*

Third, the national government informally engaged the private sector through meeting discussions to reduce COVID-19 testing prices. Fourth, financial support was provided to county governments to adapt external facilities for example hotels, schools into isolation centres in response to the pandemic.

> *“The only negotiation that may have taken place, and I’m not sure whether there was a contract, or an MOU is on the charges for testing in private sector to be reduced” National official 3, MOH*

> *“The hotel used as an isolation centre was paid for by an allocation from national government given to counties for COVID 19 services at the height of the pandemic.” County Health Accountant, County B*

The health facilities providing COVID-19 services made adaptations and invested in additional capacity to accommodate increased demand. These mainly included infrastructural changes for designated COVID-19 treatment units (CTUs), increased oxygen capacity, adaption of external private facilities and hospital spaces as isolation wards. Furthermore, there was an increase in human resource for COVID-19 services and associated capacity building activities.

> *“Initially, we did not have an ICU. There was no adequate space within the health facility. The county established an ICU in classrooms of medical training colleges.” County Chief Executive Committee member-Health, County C*

> *“We got into MOU with the private facility for quarantine for our health workers. We engaged a few private hotels where they would be accommodated. They gave us accommodation at a subsidized cost.” County Chief Officer Medical Services, County A*

#### **iii)** How to purchase

The government paid for COVID-19 services using different methods. Both national and county governments used supplementary budgets to avail resources for the COVID-19 response. The national government procured vaccines, COVID-19 supplies and paid for other COVID-19 related costs through a line-item budget. Within the study counties, the county governments paid for the waived hospital bills through a COVID-19 fund allocated from the national government. Additionally, county C reallocated funds from other departments whereas County A received in-kind donor support to enhance service provision.

> *“The county government waives isolation and treatment costs to try safeguard the residents’ welfare on out-of-pocket expenditure for those who don’t have insurance” County Director of Health Planning, County B*

> *“There was a supplementary budget that reallocated resources from different departments to COVID-19 interventions. Also, there is a national government allocation provided to support counties in the pandemic response.” County Chief Executive Committee member-Health, County C*

At national level, a conditional grant was issued from treasury through MoH to all counties to ensure the necessary investments were made for the response. Also, there were grants to cater for allowances for COVID-19 health workers and donor grants to support the country’s pandemic response (21). However, there were inadequate resources for budget allocation and county governments and health facilities experienced delays in receiving funds. To mitigate the delay, some counties set aside funds or reallocated their budgets towards the pandemic response.

> *“Resource mobilization for COVID-19 and health services in general continues to be the biggest drawback. There was an itemized 5 billion budget allocated to all counties. The counties are still independent and had their budgets for the COVID-19 response. There is the delay in funds flowing to the different counties. The COVID-19 amounts were mobilized through national treasury, coming to the ministry of health, and then released to the counties. For example, the 5 billion was received by MoH in the next financial year and then later to counties.” National official 1, MoH*

> *“In the county there was a supplementary budget that reallocated resources from different departments to COVID-19 intervention. Also, there was the national government allocation and donor partners providing support both in cash and in kind.” County Chief Executive Committee member-Health, County C*

Furthermore, health facilities experienced delays in receiving NHIF reimbursement funds due to the information technology (IT) challenges in processing NHIF claims for all services and highlighted the inadequacy of the payment methods for COVID-19 services at county level.

> *“NHIF has now become digital, the e-claim management system. We were provided with one scanner for normal NHIF clients use and the COVID-19 patients also, which could increase infection. That has led to delay and a lot of hesitance by those who are providing those services.” Medical superintendent, public hospital 1, County A*

> *“It is not sufficient because this is the same amount of money, we are using to pay our casuals. Sometimes you find that we don’t get enough supply for drugs. It is the same money we are using to buy drugs and still have to budget for other services in the hospital.” Nurse in charge, public health centre 1, County B*

NHIF enhanced schemes paid for COVID-19 testing, isolation and treatment services through the fee-for-service model. Health facilities submitted invoices for the outpatient and in-patient services offered to enhanced schemes beneficiaries. The amount was then reimbursed to the facilities.

> *“Specifically, for COVID-19 under the enhanced schemes services that have been procured for beneficiaries through NHIF, the fee for service model of payment has been applied. For outpatient services for members of enhanced schemes with limits, the fee for services usually applies” National Official 4, NHIF*

Fees-for-service payment model resulted in various reimbursement challenges experienced by NHIF. First, the lack of standardised management of COVID-19 patients across health facilities resulted in cost variation from different health providers for claims reimbursement.

> *“Initially, we didn’t have interim guidelines and standardization of management as a country. So, there was a significant cost variation across different providers because the management was more of empirical, as opposed to being informed by specific guidelines.” National Official 4, NHIF*

Second, there was increased provider-induced demand for COVID-19 in-patient admissions including ICU care. This not only increased costs of care, but also put additional pressures on the country’s critical care capacity.

> “*There was provider-induced demand, especially for admissions. When we referred to the averages some didn’t add up suggesting that some of the admissions might have been induced by the provider for clinical reasons. There were quite a few admissions to the ICU (costing 82,600 per day), yet a good number of the cases did not really require to be in the ICU. At that time the general admissions other than COVID-19 had also declined. During infection surges, this resulted in challenges in terms of access, especially for critical care.” National Official 4, NHIF*

### PFM arrangements for COVID-19 and how this influenced response to the pandemic

The national government leveraged existing public finance management (PFM) flexibilities to enhance the pandemic response. These included the establishment of a COVID-19 emergency fund (that helped to mobilise financial resources from the private sector and was established through regulations to the PFM Act published by the National Treasury), re-prioritization of planned activities, and reallocation of national and county budget amounts within shorter timelines. The PFM Act allowed for expenditure before approval. The approval for the additional expenditure was then sought within two months after the first withdrawal of the money. This process was facilitated by reallocation of resources through supplementary budgets, parliamentary approval of proposals from national treasury for supplementary budgets, and donors repurposing committed resources towards the pandemic response. This provision enabled the counties to mobilise funds to finance COVID-19 response activities without facing legal bottlenecks. The flexibilities in the budget formulation process were in accordance with the PFM Act 2012 Article 44 which allows for implementation of supplementary budgets based on fiscal responsibility and approved financial objectives (22), and the establishment of an emergency fund (22, 23) The procurement processes were adapted by allowing shorter approval timelines for expenditure of urgent commodities for the pandemic response, in accordance with the financial and procurement laws and regulations (23), and the provision of in-kind commodities from national agencies through the national government to health facilities. These measures were meant to provide additional resource mobilization and allowed timely execution of activities at the national level.

> *“The main adaptation is the COVID-19 fund which may not affect counties directly because when the monies were being drawn for the COVID-19 fund that is actually driving the response most of it went to counties directly or as in-kind.” National official 3, CoG*

> *“At national level we were able to reallocate funds and be responsive because not all the money is ring-fenced. We were meeting the timelines that had been previously set out that we wouldn’t have met had we not been in a pandemic because it wouldn’t have gotten the urgency. There has been a flurry of meetings with different agencies and departments to ensure that everything is in order but following the guidance that comes through the financing systems.” National official 1, MoH*

There was improved efficiency due to shorter timelines for procurement processes at national level. Also, there were efforts towards accountability of budgeting processes by maintaining clear documentation of received monetary amounts. However, accountability of expenditure processes experienced challenges with transparency which resulted in irregular procurement procedures at some government institutions contrary to stipulated financial procurement laws and regulations (24).

> *“There were high levels of accountability of the budgeting process by ensuring well documented planned activities and the received monetary values. This identified the gaps and the intended response to ensure that we were well prepared for the pandemic response. There was need to realign the budget and prioritize needs and to be able to make a business case for health in the long run.” National official 2, MoH*

> *“Thepositive aspect is the quick turnaround time during a period of emergency. Counties have been able to put up infrastructure (ICU beds, oxygen) in a short time which requires the adaptation to have money is flowing to the user. On the negative is the KEMSA case with PPEs. The approval of procurement processes didn’t follow the designed processes, and this led to accountability issues.” National official 1, MoH*

The NHIF financial systems were adapted to ensure timely processing of NHIF claims and were mainly responsive to the pandemic, with the exception for surveillance and auditing processes of medical claims.

> *“Provisions were made in terms of processing mechanisms to make sure that as a fund, we are not significantly affected with the revenue flows. The main issue would be in hard-to-reach areas where surveillance became quite a significant challenge. Also, the aspects of fraud management were affected because this is one of the main issues with claims where medical audits are required. The movement across to do the medical audits was affected. I wouldn’t say it’s 100% responsive.” National official 4, NHIF*

At county level, the PFM systems and processes had slight adaptations during the pandemic response. The budget formulation process and timelines remained the same. However, counties adjusted the budgets to include activities to respond to the pandemic, resulting in re-prioritization of other activities and reallocation of budgeted funds towards the pandemic response in accordance with the PFM Act 2012 Article 135 which allows for implementation of supplementary budgets upon county assembly (22).

> *“The budget development still takes the same process, from health facilities to county department, then to public participation, the county assembly approval, and finally the governor’s approval to implement the program-based budget. The only difference is most of the meetings especially the public participation and some of the assembly meetings are done virtually.” Hospital Accountant, Public Hospital 1, County A*

> *“Currently with the COVID 19, the budgeting has been affected mostly because there was need for more financing towards the pandemic” Nurse, COVID-19 centre, County B*

The flow of funds from the county government to health facilities also remained the same. County A and B health facilities generated revenue and retained the funds to spend at source upon approval by the county government. County C health facilities redirected all funds centrally to the county revenue fund (CRF). The counties found that the various aspects of the PFM systems were rigid and limited the speed and ease in which counties responded the pandemic.

> *“In terms of the PFM arrangement in counties, the processes have remained the same. For example, in terms of the monies going to the CRF, County Revenue Fund, this still happens and then funds are transferred to the special purpose account.” National official 2, MoH*

> *“The PFM structures are rigid to respond to epidemics or pandemics. This pandemic was a first and the PFM systems were not flexible for the county to respond well.” County Director of Health Planning, County B*

Across the study counties, the flow of funds had a varied effect on the pandemic response. In counties A and B, the health facilities had autonomy to spend at source and were able to procure urgent items. However, due to delays in receiving funds from the county governments, health facilities had to spend out of budget to ensure continuity of service delivery.

> *“With the Health Service Improvement Fund (HSIF), we retain funds from this financial year, we are retaining all the monies in the hospital. We budget as a hospital and we can spend” Medical superintendent, public hospital 2, County A*

> *“Most of the procurement for COVID-19 commodities were done using the allocation, the allocation by the national government. This allocation had come towards the end of the last financial year. So, during the year, by large, the funds were available.” County Health Accountant, County B*

> *“We had to review our budgets in terms of needs. Because of the delay in receiving funds, we had to spend out of budget as the managers, to be able to sustain those services and facilities” Facility in charge, public health centre 1, County A*

To mitigate the delays in fund flows affecting urgent procurement and reduce stock-outs, County A established a separate fund for the pandemic response in accordance with the Contingencies Fund and County Emergency Funds Act 2011 (25) and the PFM Regulations 2020 (23) as an additional source of funds for procurement, whereas health facilities in County B were allowed to purchase from local suppliers.

> *“The hospital had a fund which we started implementing two months before COVID-19 in 2020. So, the hospital was responsive to the pandemic. Decision making was faster. there was direct procurements, or did reverse procurements by requesting the goods, then doing the paperwork later.” Hospital Accountant, Public hospital 1, County A*

> *“There is some flexibility depending on the arising issues. The county has done local purchase ordering especially when there are delays. Also, non-pharms were purchased from local suppliers when KEMSA delayed supplies. The county purchased supplies due to COVID 19 increase and demand, like oxygen flow meters, and the pulse oximeters, and even the monitors.” Maternity nurse in-charge, public hospital 1, County B*

On the other hand, health facilities in County C redirected all revenue generated back to the CRF and health facilities were unable to urgently procure supplies and commodities required for the COVID-19 response resulting in stock outs. This was coupled by the delay in disbursement of funds from the national government with the effect trickled down to the facility level.

> *“There was a delay in disbursement of funds from the national ggovernment. Even last month the salary payment was delayed. At the lower level for facilities, this is further delayed even up to four months. This flow of resources affects the way we provide services.” County Chief Executive Committee member-Health, County C*

> *“The county sometimes relies on MOH for funds by requesting an imprest which may take very long to process. At the same time, the county may not have money, and the facilities experiences a lot of financial crunches in running day to day activities.” County Nurse In-charge, County C*

In two of the three study counties, the approval process for expenditure and procurement processes were lengthy which hindered effective procurement of urgent commodities and supplies. For instance, in county B, procurement process took up to 1 month resulting in delay of procuring urgent supplies for the pandemic response. In county C, procurement was done centrally by the county government which was associated with delays.

> *“It has been a major challenge regarding procurement of urgent supplies. Requisition process would take 1 week, and actual procurement would take 2 weeks and delivery another 1 week. For the flow of funds, the county needs approval from national government to spend their own revenue generated at the county level. The revenue must be reported to the county revenue fund first. It takes up to 3 months for counties to able to use their revenue.” County Health Accountant, County B*

> *“County C runs on a central like procurement system and with a lot of bureaucracy. The hospital is unable to meet its immediate needs, especially for large equipment. For small supplies or repairs there is a lot of delays because of the bureaucratic procurement process and contracting. Medical superintendent, public hospital 1, County C*

Contrarily, in county A, health facilities were able to procure some supplies with shorter approval timelines from the county government.

> *“Prior to COVID-19, the approval for expenditure was strict. But after that, rule was relaxed. Facilities could go back to the budget process adjust include COVID-19 supplies for example disinfectants, gloves, and then would be allowed to spend upon approval, when it’s approved you now allowed to, you are allowed to spend.” COVID-19 coordinator, County A*

The budget monitoring processes did not change as counties adhered to the rules and regulations of budget implementation to accountability especially for expenditure of funds.

> *“All those procedures were prolonged because you can’t go to the market and buy gloves and then distribute to the workers. It will become an audit query. All government entities must follow the set-out rules and regulations on how to procure goods” Hospital Administrator, public hospital 1, County B*

> *“The public finance management systems, all government entities follow the set rules and regulations, including those procurement, financial orders. They are adhered to.” Hospital administrator, public hospital 1, County C*

### COVID-19 response and financing for RMNCAH services

The funding for RMNCAH and family planning (FP) had set budgets at national level which were not affected by COVID-19 and its containment measures. However, this effect varied across the study counties. At county level, both domestic and donor funds for RMNCAH activities and procurement of commodities were reallocated for COVID-19 services. In addition, County B health facilities reallocated Linda Mama funds to finance daily operations.

> *“Most of the funds meant for RMNCAH activities, by various organizations, and even the county, were re-directed either to the COVID-19, or we were forced to program with COVID-19 in mind. It includes family planning. For example, UNFPA funding for last year was all redirected to COVID-19. Last year, the amount of money that was allocated for family planning and other Adolescent Sexual & Reproductive Health (ASRH) issues was re-diverted to COVID-19 issues” County RMNCAH coordinator, County A*

> *“The resources that were to be allocated for example, to procure reproductive health commodities were diverted to respond to the pandemic. This affected some key performance indicators like deliveries. Family planning was also affected.” County Chief Officer of Medical Services, County A*

> *“We’ve been forced to use the Linda Mama now to sustain other routine services other than the mothers themselves because the other money is being redirected. And when the hospital has no other funds other than the NHIF and the Linda Mama, we have to rely on Linda Mama.” Maternity nurse in-charge, public hospital 1, County B*

The delivery of RMNCAH and FP services was affected because of postponement of some program activities and the change in FP methods available in facilities.

> *“There has been a big change from long-term to short-term methods for family planning. This is purely because of issues of COVID-19 containment and the reduced supply of long-term methods that has led to a change in terms of how we were doing activities, including the guidelines from the ministry. Healthcare providers were advised to give short term methods over long-term methods.” RMNCAH focal person, County A*

### COVID-19 impact on health financing system and longer-term plans for UHC

The COVID-19 pandemic had both positive and negative effects on the availability of public resources for UHC. There were increased investments in the health sector and available resources at the facility level which was beneficial for all individuals seeking healthcare. The health budget allocated towards UHC was Ksh. 47.8 billion in 2019/2020 (26) and increased to Ksh. 50.3 billion in 2020/21 (27).

> *“In terms of UHC, the systems that had to be established with redirected resources from UHC supported facilities to have some certain services that they previously did not have, and they are in a slightly better position to now ensure that our primary health care systems can be rolled out without much resistance.” National official 5, development partner*

Contrarily, in County A there was re-direction of significant resources away from UHC activities to finance the pandemic response. This resulted in deprioritization of UHC related activities even at national level. In 2020/21 the national treasury allocated Ksh 1.2 billion for recruitment of additional health workers for 1 year, Ksh. 500 million to supply beds and beddings to hospitals, and Ksh 25 Million for modern walk through sanitizers to boarder points (27).

> *“As a county we had already budgeted about 20 million for the vulnerable groups like the elderly, those with chronic diseases. We were able to use that money. Primary health services were affected. We had intended to establish the community units so that we, we strengthen level one services, but that was not achieved a hundred percent.” County surveillance coordinator, County A*

> *“The UHC agenda kind of took a back seat. Some actions that may have been planned for UHC may have been delayed. There was some resource reallocation i.e., HR, finances, supplies.” National official 1, MoH*

Further reduction of financial resources for UHC was experienced with declining NHIF retention rates of formal sector employees and the statutory contributions which contribute to the UHC health benefits package.

> *“For UHC we have different pools. The formal sector employees who make statutory contributions, had a reduction in the resources available because the employment levels were affected by closure of businesses. Also, the retention rates are it’s lowest.” National official 4, NHIF*

With essential services being an integral UHC component, the study counties reallocated these resources towards COVID-19 services which resulted in interruption of essential service delivery. For instance, in County B, a gender-violence care centre was converted to the COVID-19 centre and these essential services were limited to few available rooms. In County A, health centres converted to isolation centres stopped providing essential services to communities but was reverted later. In County B, weekly specialist clinics were halted due to reallocation of staff and other resources.

> *“Yes, there was resource re-allocation, infrastructure wise, personnel, finance, supplies. The priority immediately changed to COVID-19 and it did affect other services negatively because at one point we had to stop running our special clinics.” Hospital Administrator, public hospital 1, County C*

> *“Hospital A was offering services including inpatient, outpatient, maternity laboratory services. When it was turned into an isolation facility those services were transferred elsewhere. Later, it reverted to offering all health services.” COVID-19 coordinator, County A*

Despite this, the country’s responses to the pandemic were found to be beneficial to the country’s approaches towards achieving UHC. These included: 1) the increased non-financial and financial investments in the health sector; 2) the heightened focus on preventive and primary health care which is pivotal in UHC; 3) the pandemic response affirmed the need for UHC to be prioritized; and 4) enhanced national level stakeholder engagements, coordination, and collaboration between county and national governments, further creating awareness on the importance of UHC.

> *“There was recruitment of new staff from the public service board to the counties. I believe that will go a long way in improving service delivery with the influx of patients to the facilities and other health services. Clinical Officer FBO hospital 1, County B*

> *“First, it has ended up prioritising health care. Individuals now know the need of having medical covers. Second, the preventive measures like the sanitation and washing hands has resulted in reduced expenditure on gastrointestinal diseases related to poor hygiene. Third, is the enhancement of the critical care capacity of different providers across the country.” National official 4, NHIF*

## Discussion

In this study we set out to examine how the COVID-19 pandemic and the health financing system influenced health system resilience. Our findings reveal several observations. First, the financing arrangements of COVID-19 health services varied across the different purchasers (NHIF, national and county governments). We found that COVID-19 health services were not comprehensively covered by the main purchasers of health services. For instance, the national government established a COVID-19 conditional grant for budgetary allocation to each county. This allocation enabled county governments to procure supplies and pay for COVID-19 expenses. Additionally, the national government subsidized costs for some COVID-19 supplies and commodities and purchased vaccines for provision at facility level countrywide. For isolation and treatment COVID-19 services, only NHIF enhanced schemes targeting formal sector employees paid for these services. This implied that uninsured individuals and majority of the population in general NHIF schemes incurred out of pocket expenses for high cost COVID-19 services. As of 2019, the NHIF coverage was 18% of Kenyans leaving majority of the population uninsured(28). Over 80% of the Kenyan workforce are in the informal sector, and majority of them are either not eligible or cannot afford the premiums set by the government to maintain health insurance provision (29). With the livelihoods of individuals threatened due to the negative socio-economic impact of the pandemic, this further created a financial barrier and inequity of access to COVID-19 services for the indigent and vulnerable populations.

Second, there was partial flexibility of PFM systems and processes to enable the health system respond to the pandemic. At national level, there was re-prioritization of activities and reallocation of budgeted funds towards the pandemic response. This was coupled with shorter timelines for procurement of supplies. In emergency events like pandemics, the budget space should allow for resources to be moved from other sectors and programmes to support the health sector, and the process of realignment is much easier when budgets are designed along programme lines (30). However, the county governments maintained the PFM structures and systems to ensure accountability and fiscal responsibility as stipulated the by PFM Act 2012. This resulted in challenges to efficiently respond to the pandemic, as characterised by prolonged timelines for flow of funds from the national to county governments and to health facilities, and lengthy procurement and approval for expenditure processes at county level. Counties were unable to procure urgent items to effectively provide services leading to stock-outs and affecting service delivery. On the positive, two of the three study counties were able to reallocate resources through supplementary budgets and finance COVID-19 infrastructure and approve procurement within shorter timelines. The resilience of health systems may be determined by quickly adjusted PFM rules and procedures, and fiscal arrangements such as direct budget transfers that aim to accelerate release of funds (8).

Third, the reallocation of various resources (financial, human resources, infrastructure) towards the pandemic response negatively impacted some essential health services. We found that financing of RMNCAH services and the impact on service delivery varied. Two out of the three counties reprioritized and reallocated domestic and donor funds for RMNCAH program activities and procurement of FP supplies to finance COVID-19 services. This resulted in postponement of program activities and change in the FP methods utilized in health facilities which could affect FP choice and uptake. Additionally, Linda Mama funds were utilized for operational costs where alternative funding sources lacked. For infrastructure, health centres used as isolation centres were required to stop providing essential health services for the period. Consistent with our findings, similar settings reported a downward trend in specific outpatient services due to COVID-19 (31–33). This disruption of essential service delivery provides warning signs of the negative effect of years of progress made to improve reproductive and essential health indicators.

Fourth, the impact of COVID-19 health financing on the availability of public resources for UHC was found to be bi-directional. At the county and national level, the implementation of UHC related activities slowed down due to re-prioritization and re-allocation of funds towards the pandemic response. With NHIF as the main driver for financing UHC, there was reduced financial resources to support the UHC health benefits package, and this was attributed to decreased membership and statutory contributions due to job losses Additionally, COVID-19 resources were not directly transferred to health facilities, as this would be an avenue to ensure health system resilience. This highlights the negative effects of the pandemic response towards the progress of UHC in Kenya. On the positive, the intensive resource mobilization for the health sector to respond to the pandemic strengthened areas that were lacking prior (infrastructure, human resource), and this was beneficial to all individuals seeking healthcare services. The focus of these resources on preventive health measures like handwashing and sanitation, contact tracing amongst others, contribute to strengthening primary health care which is a fundamental component of achieving UHC (34). Additionally, national level multisectoral coordination experienced in the COVID-19 response builds awareness and advocacy on the importance of universal access to healthcare and the need for adequate investment to strengthen health system inputs to achieve this goal (35).

A key limitation of the study was the small number of study counties as the health financing arrangements varied across the counties. Although our findings may not be generalizable, we unpack the issue of interest within the context of national and county levels which can be analytically generalizable. Notwithstanding, the main strength of the study is the qualitative inquiry which targeted interviews across all levels of health sectors, providing an understanding of the impact of COVID-19 health financing from both public and private sectors, and at the levels of policy formulation to implementation of the pandemic emergency response. Several recommendations can be drawn from our findings. First, these findings underscore the need for the NHIF to harmonize its benefit packages to enhance equity. While inequitable entitlements have characterized NHIF schemes before COVID-19, the actions taken by the NHIF during the pandemic to pay for COVID-19 services under the enhanced scheme only have amplified these inequities. Second, there is a need for a systematic process for defining service entitlements generally, and during pandemics specifically. Third, the national and county governments should develop formal mechanisms for engaging the private sector to enhance surge capacity during emergencies. Fourth, while PFM flexibilities exist to enhance country response to pandemics, there is a need for a review of the processes for activating emergency funds to eliminate bureaucratic bottlenecks that lead to delays in their activation. Lastly, the government should strengthen accountability mechanisms for PFM flexibilities during pandemics. Specifically, this includes strengthening accountability for emergency procurement and budget expenditures during health emergencies.

## Conclusion

For the past three years, the COVID-19 pandemic has tested the ability of the Kenyan health system to withstand crises while maintaining routine functions. This has highlighted the strengths and weaknesses of the health financing functions that have influenced the adaptability, responsiveness, and capacity of health system’s response to the pandemic. Strengthening health systems to improve their resilience to cope with public health emergencies requires substantial investment to strengthen financing systems and a legal framework that allows PFM flexibility (national and county levels) to respond to health emergencies. Health financing arrangements are integral in determining the extent of adaptability, flexibility, and responsiveness of health system to COVID-19 and future pandemics.

## Data Availability

The datasets generated and/or analysed during the current study are not publicly available due to participant confidentiality but are available from the corresponding author [AK] on reasonable request.

## Supporting information

S1 Appendix Ethical approval for the study

## Acknowledgements

We acknowledge the support of the county health departments and health facilities that participated in this study; Felix Murira, Shano Guyo, and Daniel Koech from ThinkWell Kenya.

